# Examining public acceptance of AI versus human-centric dementia care across NHS England’s dementia pathway stages

**DOI:** 10.1101/2025.10.29.25339118

**Authors:** Thomas O’Fee, Santosh Vijaykumar, Michael Craig

**Affiliations:** Climate & Health Infodemic Resilience Programme (CHIRP), Northumbria University, Newcastle upon Tyne, United Kingdom; Psychology and Communication Technology (PaCT) Lab, School of Psychology, Faculty of Health and Wellbeing, Northumbria University, Newcastle upon Tyne, United Kingdom; School of Psychology, Faculty of Health and Wellbeing, Northumbria University, Newcastle upon Tyne, United Kingdom

**Keywords:** Artificial Intelligence (AI), Dementia Care, Smart Home Technology, Acceptability, User Agency

## Abstract

With dementia diagnoses in the UK projected to exceed one million in 2025, there is an urgent need for scalable and effective care solutions to ease pressure on health and social care systems. Artificial Intelligence (AI)-enabled smart home systems are emerging as promising digital health innovations, offering cognitive support, real-time monitoring, and decision-making assistance. However, concerns around trust, user agency, and misconceptions about AI continue to limit acceptance and may hinder adoption. This study examined how stage of dementia care, system design, and level of system involvement shape public attitudes towards AI-driven care technologies. A repeated-measures design was employed, using vignettes that varied across three dimensions: stage of care (aligned with NHS England’s Dementia Well Pathway), system centrism (AI-based versus human-based), and level of involvement (moderate support versus full control). Participants aged 55-64 years – a group at elevated risk of developing dementia or encountering these technologies within the next decade – rated each scenario on both acceptability and likelihood of use. Findings showed that acceptability was sensitive to both care stage and system type. A significant interaction indicated that full human involvement was consistently rated as more acceptable, while AI involvement was viewed more favourably only at moderate levels. This interaction effect intensified across the dementia care pathway, with the largest discrepancies observed in later stages such as Living well and Dying well, where full-control AI was rated least acceptable. In terms of perceived likelihood, scenarios were generally judged more likely under conditions of moderate involvement, with human-centric scenarios rated as more likely than AI-centric ones, particularly in later stages. These results highlight the importance of trust, autonomy, and public understanding in AI adoption. Acceptance was highest when AI was positioned to augment rather than replace human input, supporting hybrid models that preserve agency while enhancing scalability.

**Author Summary:** Dementia diagnoses are increasing rapidly, creating an urgent need for innovative care solutions. Artificial Intelligence (AI) and AI-integrated smart-homes offer a promising way to provide cognitive care and support through remote monitoring, but public trust and acceptance remain a challenge. Here, we explored people’s affective attitudes towards AI for dementia care by presenting scenarios that varied based on stage of dementia (from preventative to end-of-life care), whether the system was AI or human-based, and the level of control the system had over the care (moderate or full). We surveyed a sample of older adults, a group of potential future users of this technology. Our findings show that people’s attitudes are highly dependent on factors such as stage of care, level of system control/involvement, and system centricity, human-centric systems were consistently rated as the most acceptable option, while AI was most favourable when offering moderate support rather than taking full control. This preference for human-centric, moderate support grew stronger in the later stages of dementia care. In summary, people are more open to adopting AI when it is used to enhance human care, rather than replace it. This suggests that hybrid healthcare models, where AI is used to support and supplement caregivers, will be the most effective way to integrate these scalable technologies into dementia care.

## Introduction

In 2025, it is projected that the number of people with a clinical diagnosis of dementia will surpass one million, making it increasingly likely that almost everyone will know someone affected by the condition [1]. Dementia is not a single disease, but an umbrella term used to describe a group of neurodegenerative conditions, such as Alzheimer’s disease, which impair memory and other cognitive abilities and behaviours that interfere significantly with a person’s ability to maintain activities of daily living [2]. Despite the profound challenges posed by dementia, older adults express a strong desire to live independently and maintain autonomy for as long as possible [3]. However, the later stages of neurocognitive disorders are characterised by behavioural disturbances and psychological symptoms, necessitating increased levels of professional care and living support and, often, a loss of sense-of-self [4].

The prevalence of this condition and the growing care demand have prompted extensive research aimed at developing effective treatments and enhancing support networks [5]. Global initiatives such as The UN Decade of Healthy Ageing underscore the urgency of not only extending life expectancy but also ensuring that additional years are accompanied by good quality of life [6]. This initiative has catalysed the investigation of innovative long-term care solutions designed to reduce the burden on unpaid carers and professional care staff [7]. Among these solutions, emerging technologies such as Artificial Intelligence (AI) are increasingly recognised for their potential to enhance autonomy, support daily functioning, and improve the safety of older adults within their homes.

Human-Computer Interaction (HCI) researchers have increasingly focused on the integration of AI in Gerontechnology and dementia care, designing intelligent systems that can understand, predict, and respond to the unique and dynamic needs of users [8]. Advances in this area are widely reported in the media, often in ways that anthropomorphise or demonise AI systems, contributing to public concern regarding ethical deployment, agency, and the role of AI in sensitive healthcare contexts [9]. Nonetheless, AI and machine learning have demonstrated clear benefits in healthcare, from early detection of cognitive decline to supporting patient management and care coordination [10, 11]. More recently, AI-driven systems have been implemented in home environments through Internet of Things (IoT) devices, integrating sensors and telehealth systems to create safer, more supportive spaces for older adults [12, 13, 14].

These AI-integrated home care systems offer a wide range of potential benefits. They can provide prospective memory support [8], monitor patient behaviours and generate reports for caregivers [15, 16], and detect early signs of cognitive decline using task performance assessments [17]. In addition, AI can enable personalised preventative measures and interventions, potentially slowing functional decline and enhancing quality of life [18]. As the market for these technologies expands, older adults are increasingly recognised as a key consumer group [19]. However, adoption remains limited, in part due to reluctance to integrate AI into health and cognitive care and lack of familiarity with the technologies [20]. Research indicates that people often prefer human over automated healthcare, even when the AI may deliver objectively equivalent care, suggesting a gap between perceived and actual utility [21].

One factor that may influence adoption is the perception of agency: if a system is perceived to limit personal control, it may be considered less acceptable regardless of its practical benefits. Lay beliefs about AI, including assumptions about its autonomy, accuracy, and decision-making capabilities, act as heuristics that influence user trust and willingness to adopt AI in complex health contexts [22, 23]. Misconceptions about AI, particularly around themes of involvement, agency, and reliability, may therefore significantly shape attitudes toward AI-integrated smart home systems, affecting the perceived appropriateness of these technologies at different stages of care. Furthermore, theoretical models of technology acceptance, such as the Technology Acceptance Model (TAM) and Unified Theory of Acceptance and Use of Technology (UTAUT), suggest that perceived usefulness and perceived ease of use are not static but context dependent [24]. In the context of dementia care, the stage of illness may moderate acceptability because the perceived risks and benefits of relinquishing control to a system are likely to vary with levels of dependency: for example, early-stage users may prioritise autonomy and independence, whereas later-stage users may prioritise safety and support, influencing their tolerance for highly involved AI interventions.

Building on this evidence, the present study aims to investigate how perceived system involvement impacts the willingness of older adults to integrate dementia-focused smart home systems driven by AI. Specifically, the study addresses two key questions: (1) Does the type of system (AI versus human) influence user trust and acceptability more than perceived loss of agency? and (2) Does the stage of care influence which systems are perceived as most beneficial? These questions are grounded in evidence that acceptance of healthcare technologies is context-dependent, shaped by both the perceived capabilities of the system and the stage of illness or dependency of the patient.

We hypothesised that AI systems would be perceived as less acceptable than human-centric systems across all stages of care. We further hypothesised that this difference would be more pronounced at later stages, such as the “living” and “dying” stages, when users may be most sensitive to potential losses of autonomy. We also predict that highly involved AI systems, those that take a more active role in care, will be perceived as less acceptable than less-involved AI systems, due to concerns about diminished agency. By employing a vignette-based survey methodology, this study will examine attitudes toward both AI and human systems, providing insight into how involvement and stage of care interact to shape perceptions of acceptability and trust in AI-integrated dementia care.

## Methods

### Ethics statement

This research was approved by the Faculty of Health and Wellbeing Research Ethics Committee at Northumbria University (Ref: 7477). Informed online consent was acquired from all participants following an initial study briefing and procedures adhered to the ethical principles for research in humans.

### Design

We employed a repeated measures design that used within-subject factors. There were 3 factors used as independent measures; *Stage* refers to the stage of the NHS Dementia-Well pathway that the scenario was depicting, this could be “Preventing-well”, “Diagnosing-well”, “Supporting-well”, “Living-well”, or “Dying-well”. *Centrism* refers to the basis of the system being depicted in the scenario, an AI-based system, or a human-based (no AI) based system. Finally, *Level of Involvement (LoI)* refers to the role played by the system, “moderate” indicates a supportive, yet collaborative role providing supplementary aid where some aspects of care are also managed by human oversight (in AI-centric scenarios) or personal agency (in human-centric scenarios), whereas “full” depicts the system having total control over decisions and actions related to the dementia care. Participants completed the online study, which was delivered via the Qualtrics research platform, on their personal laptop or PC device. The study was completed in a single session lasting approximately 30 minutes.

### Participants

Power analyses were conducted using G*Power 3.151 [25] to determine the minimum sample size required to detect main effects, interactions, and pairwise comparisons in the 5 (Stage) × 2 (Centrism) × 2 (Level of Involvement) repeated measures design. A medium effect size (f = 0.20) and power of 0.80 were specified for all analyses. This was based on mixed evidence to support the use of either medium or large effects from previous literature. For example, Lennartz [26] found a range of effect sizes when investigating attitudes towards levels of AI control in diagnosis and treatment planning (d = 1.68) and for different disease severity (d = 0.426). For main effects and interactions, an alpha level of 0.05 was used. For pairwise comparisons, an adjusted alpha level of 0.005 was applied to control for multiple comparisons. The largest required sample size was observed for pairwise comparisons, which indicated a minimum of 58 participants. This sample size ensures sufficient power to detect the hypothesised effects across the study’s factors. This was exceeded through the recruitment of 76 adults aged 55-64 years old (M age = 59.00, SD = 3.00; 33 females, 43 males). Our sample consisted of 90.7% English, Welsh, Scottish, Northern Irish or British Ethnicity, with the remaining 9.3% consisting of other White backgrounds, Black, Arican or Caribbean backgrounds, Indian, Bangladeshi, White and Asian, or other Asia backgrounds equally.

All participants were recruited through Prolific.com, an online research platform where participants are verified and vetted. Inclusion criteria required participants to live in England (United Kingdom) and be between 55-64 years old. The age criterion was used because dementia typically occurs in individuals aged 65+ and thus 55-64 years old individuals are high-risk and the next generation to possibly develop dementia [27], which makes their attitudes towards novel dementia interventions especially relevant. Participants were not eligible if they had a diagnosed neurocognitive disorder or known problems with thinking skills. These individuals were recruited between 22^nd^ to 23^rd^ October 2024 through Prolific’s participant panel and reimbursed £2.50 for their time.

### Materials

Participants in the study completed pre-test measures. The Dementia Knowledge Assessment Tool (DKAT) [28] was used to measure individuals’ knowledge about dementia by presenting statements about the condition that participants would identify as true or false. The Dementia Attitude Scale [29], a 20-item, Likert scale questionnaire, was used to measure participants’ attitude towards dementia, assessing their knowledge about the condition and their comfort towards interacting with a diagnosed patient. The Artificial Intelligence Literacy Scale [30] was used to measure participants’ understanding of AI as well as their ability to utilise it. Conditions were in the form of AI-generated, hypothetical vignettes. Each vignette was created with a specific set of instructions and would depict fictional scenarios that revolved around how we detect and care for people with dementia. For outcome measures, the Theoretical Framework for Acceptability (TFA construct) [31] was used as to assess the perceived acceptability of a given intervention, a framework used to assess the design and implementation of healthcare interventions considering factors such as self-efficacy, ethicality, affect, and perceived effectiveness. Likelihood was another outcome measure, meaning participants rated how likely they thought the scenario was to become reality. This was collected using a single Likert scale question within each condition.

### Procedure

Data were collected via an online survey administered through the Qualtrics research platform. Firstly, participants were requested to read a participant information sheet and complete an informed consent sheet. If consent was provided, they proceeded to complete a set of demographic questions, where data on their age, gender, highest level of education, ethnicity, current employment, and relationship with anyone who has experienced dementia were collected. Following this, they completed three short surveys that probed their knowledge of dementia via the Dementia Knowledge Assessment Tool [28], awareness of dementia via the Dementia Attitude Scale [29], and comprehension of artificial intelligence via the Artificial Intelligence Literacy Scale [30].

Beginning the task, participants would be prompted to read one vignette at a time, recording responses to both outcome measures after each one. Each vignette would depict a fictitious scenario where the subject of the story is engaging with a care system, whether it be AI or Human, and moderate involvement or full involvement. It would also depict a specific stage in the NHS Dementia-well Pathway; Preventing, Diagnosing, Supporting, Living, or Dying. Participants would, for each vignette, fill out the TFA questionnaire, probing attitudes towards comfort, effort, fairness, confidence, efficacy and acceptability, followed by a single question asking them how likely they think the scenario is to become reality, which read “How likely do you think this system is to become reality”. Once all questions were answered, they could proceed to the next scenario. There were a total of 20 fictitious scenarios. All study materials, including vignettes, will be made available on the project OSF site at https://osf.io/wautg/.

#### Example Vignette

Diagnosing-Well Stage, AI-centric, Moderate-involvement

*“David uses a smart home device that tracks his cognitive function through daily activities. The AI system flags potential signs of cognitive decline and recommends that he visit his GP for further testing. While helpful, the AI acknowledges that its findings are preliminary and must be confirmed by a medical professional. This collaborative approach ensures that David receives an accurate diagnosis and appropriate care, combining AI assistance with expert medical evaluation. The AI provides useful insights and reminders, but the final diagnosis and treatment plan are developed by his healthcare team.”*

Following completion of all conditions, participants were presented with a debrief sheet that provided detailed information about the study purpose and thanked for their time.

### Data Analysis

Statistical analyses were conducted in R, an open-source and industry standard programming language [32]. Descriptive statistics were computed for participant demographics and performance across our dependent measures, acceptability and likelihood, as well as for the DKAT, DAS, and AILS. The mean values for the TFA construct, and the raw Likelihood rating scores were used as dependent variables during the analyses. Repeated measures analyses of variance (RMANOVAs) were used to examine main effects of within-subject factors: centrism (AI vs. human), level of involvement (moderate vs. full), and NHS England’s dementia-well pathway (preventing-well vs. diagnosing-well vs. supporting-well vs. living-well vs. dying-well), and interactions between these within-subject factors. Any significant interaction effects were followed up with pairwise comparisons and corrected for multiple hypotheses using Bonferroni corrections.

## Results

### Effect of Centrism and Level of Involvement on Acceptability

A three-way repeated measures ANOVA was conducted to examine the effects of Dementia-Well pathway Stage, Centrism (AI vs Human-centric), and level of involvement (Moderate vs Full) on the ratings of perceived acceptability. Tests were conducted using logarithmically transformed data due to abnormal data distributions. This revealed a significant main effect of level of involvement, *F*(1,75) = 68.54, *p* < .001, n^2^_p_ = .54, 95%CI [0.38, 0.65] because, overall, participants rated full involvement (M = 3.75, SD = 0.93) as more acceptable than moderate involvement (M = 3.31, SD = 0.95). No significant main effect of centrism was observed (*F*(1,75) = .26, *p* =.611), meaning there was no significant difference in perceived acceptability between human-centric scenarios (M = 3.54, SD = 1.05) were rated and AI-centric scenarios (M = 3.52, SD = 0.87) as a whole. Lastly, a significant main effect of pathway stage, *F*(3.57, 268.02) = 38.38, *p* < .001, n^2^_p_ = .27, 95%CI [0.18, 0.34], was revealed. This was due to ratings of acceptability trending towards being lower for later stages of the dementia pathway.

Regarding interactions, significant interactions were observed between level of involvement and centrism (*F*(1, 75) = 389.54, *p* < .001, n^2^_p_ = .86, 95%CI [0.80, 0.89]), level of involvement and stage, *F*(3.67, 275.41) = 43.23, *p* <.001, n^2^_p_ = .39, 95%CI [0.31, 0.46], and centrism and stage, *F*(3.70, 277.39) = 6.35, *p* < .001, n^2^_p_ = .05, 95%CI [0.01, 0.10]. A significant three-way interaction between level of involvement, centrism, and stage was also observed, *F*(3.53, 264.84) = 64.05, *p* < .001, n^2^_p_ = .44, 95%CI [0.36, 0.51]. Follow up pairwise tests with a Bonferroni correction indicate that across all stages, there was a significant difference between the acceptability of full AI (M = 3.22, SD = .92) compared to moderate human (M = 2.80, SD = .89) systems (*t*(75) = 4.258, *p* < .001) and a significant difference between full human (M = 4.28, SD = .57) and moderate AI (M = 3.82, SD = .71) systems (*t*(75) = -7.610, *p* < .001).

To explore the significant interactions further, a series of five follow-on two-way ANOVAs using within-subject factors level of involvement (moderate vs. full) and centrism (human vs. AI) were conducted, one for each of the dementia pathway stages. Figure 1 shows mean scores and interaction effects for each of the dementia pathway stages.

**Fig 1.**
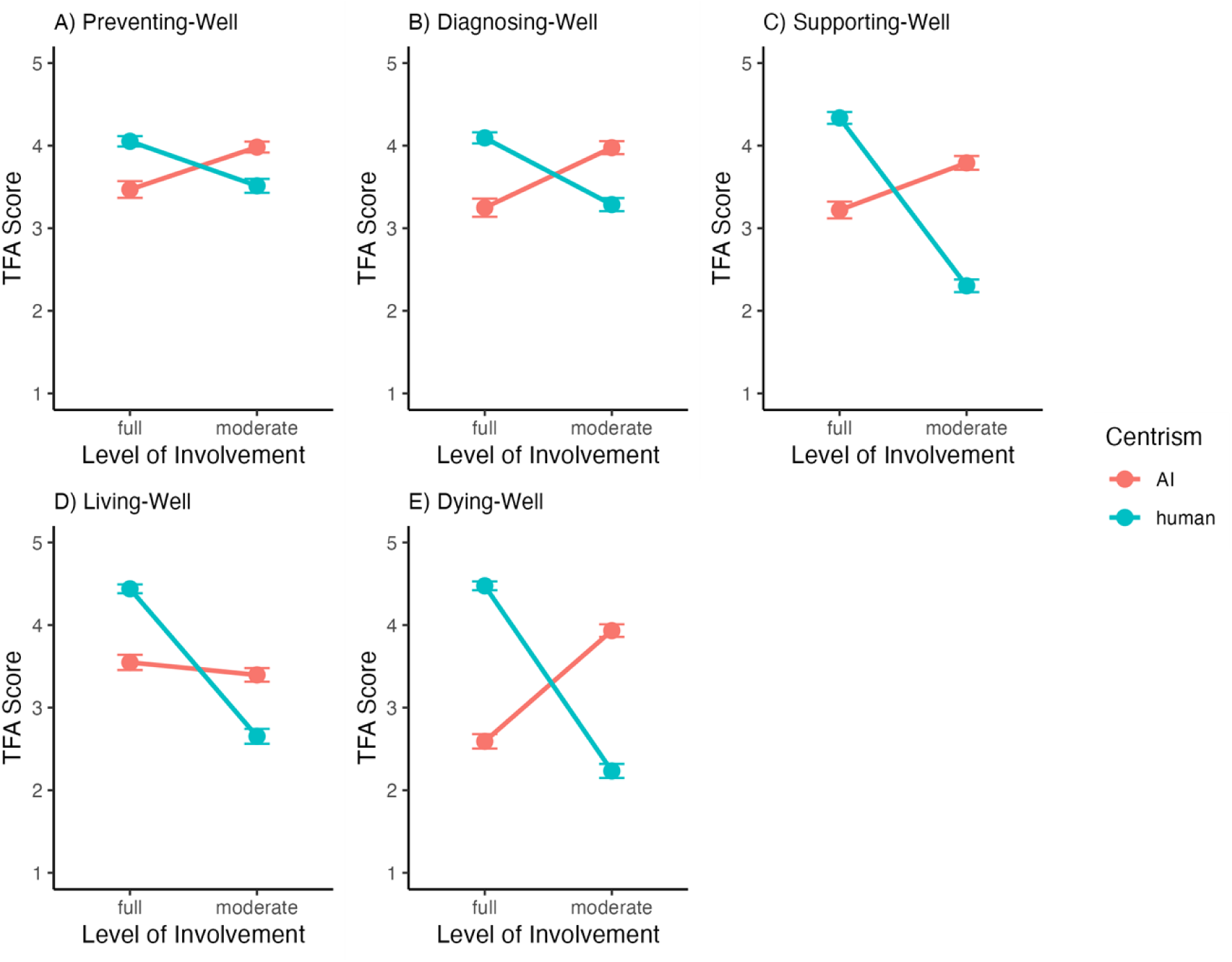
Ratings of Acceptability by pathway stage. **Note.** This figure presents participants’ ratings of acceptability for AI-centric (red lines) versus human-centric (blue lines) systems with varying levels of involvement across the Dementia-Well pathway stages. Acceptability depends on the interplay between care stage, system centrism, and level of involvement. Early stages (Preventing-well and Diagnosing-well) show more openness to AI, even at higher involvement levels, whereas later stages (Supporting-well, Living-well, and Dying-well) indicate strong preference for fully involved human-centric systems. Moderately involved AI systems remain relatively acceptable across all stages, highlighting their potential as supportive, collaborative tools.

#### Preventing-Well

There were no significant main effects of Centrism, (F(1,75) = 0.71, *p =* .401) or LoI, *(F(*1, 75) = .02, *p* = .880) (see Fig 1a). Meaning that there were no significant differences between AI (M = 3.73, SD = 0.78), or Human (M = 3.79, SD = 0.70) systems, nor any differences between Full (M = 3.70, SD = 0.78) or Moderate (M = 3.75, SD = 0.70) involvement. However, there was a significant interaction effect, F(1, 75) = 56.76, *p* < .001, n^2^_p_ = 0.45, 95% CI[0.29, 0.58] which indicated that an effect of Centrism was dependent on the level of involvement of the system in the scenario. However, a follow up pairwise comparison revealed no significant difference between Full-AI (M = 3.47, SD = 0.88) and Moderate-Human systems (M = 3.51, SD = 0.73), (*t*(75) = .393, *p* = 1.00), nor any significant difference between Moderate-AI (M = 3.98, SD = 0.58) and Full-Human (M = 4.05, SD = 0.55) systems, (*t*(75) = - 0.825, *p* = 1.00).

#### Diagnosing-Well

At the “Diagnosing-well” stage (Fig 1b), a two-way ANOVA with Centrism and LoI was conducted with TFA score as the dependent variable. Results revealed no main effects of Centrism or LoI, (F(1, 75) = 1.94, *p* = .168, F(1, 75) = .00, *p* = .992). Meaning no significant differences between AI (M = 3.61, SD = 0.90) and Human (M = 3.69, SD = 0.75) conditions, nor between fully involved (M = 3.67, SD = 0.90) or moderately involved (M = 3.63, SD = 0.77) conditions. However, there was a significant interaction effect between Centrism and LoI, F(1, 75) = 86.65, *p* < .001, n^2^_p_ = 0.54, suggesting that a large effect only exists under specific conditions regarding whether the system was AI or Human centric, and whether it depicted Moderate or Full involvement. Pairwise comparisons reveal no significant difference between Full-Human (M = 4.09, SD = 0.59) and Moderate-AI (M = 3.98, SD = 0.68) conditions, (*t*(75) = 1.364, *p* = 1.00), or Full-AI (M = 3.25, SD = 0.96) and Moderate-Human (M = 3.29, SD = 0.69) conditions, (t(75) = -0.288, *p* = 1.00).

#### Supporting-Well

At the “Supporting-well” stage (Fig 1c), the two-way ANOVA revealed a significant main effect for Centrism, (*F*(1, 75) = 9.77, *p* = .003, n^2^_p_ = 0.12), and a significant main effect for LoI, *F*(1, 75) = 82.61, *p* < .001, n^2^_p_ = 0.52). Meaning that there was a significant difference in perceived acceptability between AI (M = 3.50, SD = 0.86) and Human (M = 3.32, SD = 1.21) conditions and between Fully involved (M = 3.78, SD, = 0.95) and moderately involved (M = 3.05, SD = 1.02) conditions. There was also a significant interaction effect, *F*(1, 75) = 267.40, *p* < .001, n^2^_p_ = 0.78 suggesting that an effect only exists under specific conditions regarding whether the system was AI/Human centric, and whether it depicted Moderate or Full involvement. Follow-up pairwise comparisons revealed that there was a significant difference between Full-AI (M = 3.22, SD = 0.88) and Moderate-Human (M = 2.30, SD = 0.66) conditions, *t*(75) = 7.259, *p* < .001 and a significant difference between Moderate-AI (M = 3.79, SD = 0.73) and Full-Human (M = 4.34, SD = 0.62) conditions, *t*(75) = -5.442, *p* <.001.

#### Living-Well

At the “Living-well” stage (Fig 1d), a two-way ANOVA with Centrism and LoI was conducted with TFA score as the dependent variable. Results revealed a significant main effect for LoI, *F*(1, 75) = 126.39, *p* < .001, n^2^_p_ = 0.63, indicating a significant difference between Full (M = 3.99, SD = 0.79) and Moderate (M = 3.02, SD = 0.84) conditions (see Figure 1d). However, there was no significant effect of Centrism *(F*(1, 75) = 0.11, *p* = .737). There was also a significant interaction between Centrism and LoI on TFA score, *F*(1,75) = 96.35, p < .001, n^2^_p_ = 0.56. Follow-up pairwise comparisons (Bonferroni) indicate significant differences between both Full-AI (M = 3.56, SD = 0.81) and Moderate-Human (M = 2.65, SD = 0.79) conditions *(t*(75) = 6.851, *p* <.001), as well as between Moderate-AI (M = 3.40, SD = 0.72) and Full-Human (M = 4.44 , SD = 0.46) conditions, (*t*(75) = -10.197, *p* < .001) (see Figure 1d).

#### Dying-Well

At the “Dying-well” stage (Fig 1e), the two-way ANOVA for Centrism and LoI on TFA score revealed a significant main effect of LoI, *F*(1, 75) = 33.70, *p* < .001, n^2^_p_ = 0.31, meaning there was a significant difference between acceptability for Full (M = 3.53, SD = 1.14) conditions and for Moderate (M = 3.08, SD = 1.10) conditions There was no significant main effect of Centrism, *F*(1,75) = .09, *p* = .762, however, there was a significant large interaction effect, *F*(1, 75) = 395.46, *p* <.001, n^2^_p_ = 0.84. Follow-up pairwise comparisons (Bonferroni) revealed a significant difference between Full-AI (M = 2.59, SD = 0.76) and Moderate-Human (M = 2.23, SD = 0.74) conditions (*t*(75) = 2.88, *p* = .031), as well as between Moderate-AI (M = 3.93, SD = 0.67) and Full-Human (M = 4.48, SD = 0.47) conditions (t(75) = -6.747, *p* < .001).

### Effect of Centrism and Level of Involvement on Likelihood

A three-way repeated measures ANOVA was conducted to examine the effects of Dementia-Well pathway Stage, Centrism (AI vs Human-centric), and level of involvement (Moderate vs Full) on the ratings of perceived likelihood. This revealed a significant main effect of Level of Involvement, *F*(1,75) = 92.76, *p* <.001, n^2^_p_ = .58, 95%CI [0.44, 0.69] because, overall, participants rated Full Involvement (M = 3.17, SD = 1.28) as more likely than Moderate Involvement (M = 4.24, SD = 0.79). There was also a significant main effect of Centrism was observed (*F*(1,75) = 21.84, *p* < .001, n^2^_p_ = 0.30), meaning there was a significant difference in perceived likelihood between Human-centric scenarios (M = 3.89, SD = 1.19) were rated and AI-centric scenarios (M = 3.51, SD = 1.17) as a whole. Lastly, a significant main effect of Pathway Stage, *F*(3.67, 274.91) = 5.02, *p* < .001, n^2^_p_ = .05, 95%CI [0.01, 0.10], was revealed.

Regarding interactions, a significant interaction was observed between Centrism and Pathway Stage (*F*(3.39, 254.38) = 6.74, *p* < .001, n^2^_p_ = .10, 95%CI [0.04, 0.16]), but no significant interactions between level of involvement and stage (*F*(3.69, 276.68) = 2.29, *p* = .066), or Involvement and Centrism, (*F*(1, 75) = 2.14, *p* = .147).However, A significant three-way interaction between Level of Involvement, Centrism, and Stage was observed, *F*(3.62, 271.75) = 3.46, *p* = .010, n^2^_p_ = .04, 95%CI [0.00, 0.08]. Follow up pairwise tests with a Bonferroni correction indicate that across all stages, there was a significant difference between the acceptability of Full-AI (M = 2.96, SD = 1.26) compared to Moderate-Human (M = 4.41, SD = 0.81) systems, (*t*(75) = -10.524, *p* <.001) and a significant difference between Moderate-AI (M = 4.08, SD = 0.74) and Full-Human (M = 3.38, SD = 1.28) systems (*t*(75) = 6.413, *p* < .001).

To explore the significant interactions further, a series of five follow-on two-way ANOVAs using within-subject factors Level of Involvement (Moderate vs. Full) and Centrism (Human vs. AI) were conducted, one for each of the dementia pathway stages.

#### Preventing-Well

At the Preventing-Well stage (see Fig 2a), a 2-way ANOVA revealed a significant main effect of Level of Involvement, *F*(1, 75) = 60.15, *p* < .001, n^2^_p_ = .47, 95% CI [0.31,0.60], where scenarios depicting Moderate involvement (M = 4.28, SD = 0.63) were considered significantly more likely than scenarios depicting Full system involvement (M = 3.24, SD = 1.22). However, there was no significant main effect of centrism (*F*(1, 75) = .68, *p* = .411), meaning there was no significant difference in likelihood between AI (M = 3.78, SD = 1.02) and Human (M = 3.73, SD = 1.18) conditions. Nor was there a significant interaction effect (*F*(1, 75) = 0.65, *p* = .467).

**Fig 2.**
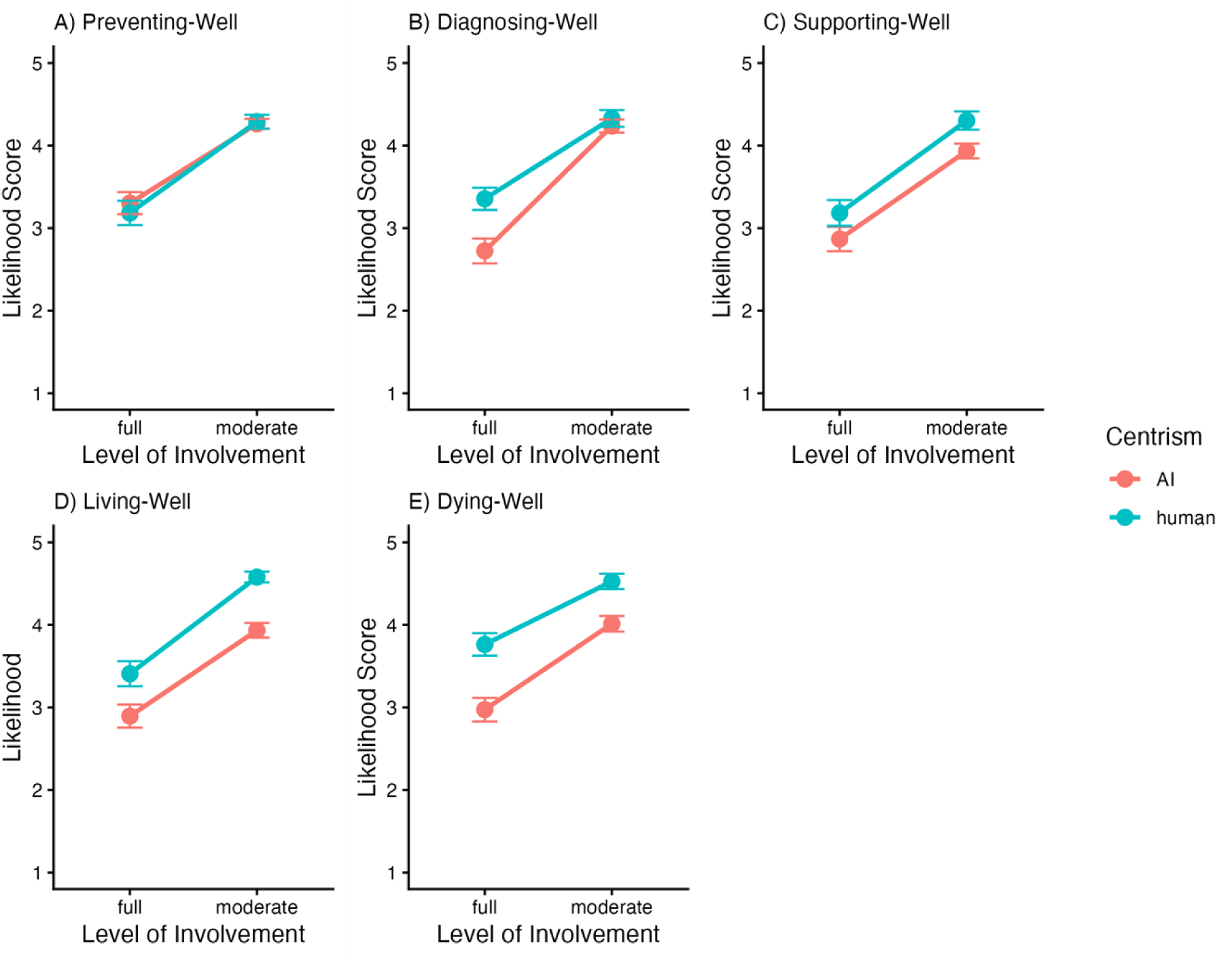
Ratings of likelihood across the pathway stages. **Note.** This figure illustrates participants’ ratings of the likelihood of AI-centric (red lines) versus human-centric (blue lines) systems with varying levels of involvement across the Dementia-Well pathway stages. Consistently, human-centric systems with moderate involvement were perceived as the most probable scenario, particularly in later stages, while fully involved AI systems were generally viewed as less likely.

#### Diagnosing-Well

At the Diagnosing-Well stage (Fig 2b), the 2-way ANOVA revealed significant main effects of Level of Involvement, *F*(1, 75) = 67.42, *p* < .001, n^2^_p_ = 0.47, 95% CI [0.31, 0.60], and Centrism, *F*(1, 75) = 12.80, *p* <. 001, n^2^_p_ = 0.15, 95% CI [0.03, 0.30]. This indicates significant differences between both Full (M = 3.04, SD = 1.28) and Moderate (M = 4.28, SD = 0.79) conditions, and between AI (M = 3.48, SD = 1.29) and Human (M = 3.84, SD = 1.15) conditions. There was also a significant interaction effect, *F*(1, 75) = 13.35, *p* < .001, n^2^_p_ = 0.15, 95% CI [0.03, 0.30], which in follow-up pairwise comparisons indicate both significant differences between Full-AI (M = 2.72, SD = 1.31) and Moderate-Human (M = 4.33, SD = 0.89) conditions (*t*(75 = -7.853, *p* < .001), as well as between Moderate-AI (M = 4.24, SD = 0.69) and Full-Human (M = 3.36, SD = 1.17) conditions (*t*(75) = 6.592, *p* < .001)

#### Supporting-Well

At the Supporting-Well stage (Fig 2c), there were significant main effects of Level of Involvement, *F*(1,75) = 51.88, *p* <.001, n^2^_p_ = 0.42 95%CI [0.24, 0.54], and of Centrism, *F*(1,75) = 4.33, *p* = .041, n^2^_p_ = .05, 95%CI [0.01, 0.18]. This indicated that human-centric scenarios (M = 3.74, SD = 1.30) were rated as more likely than AI-centric (M = 3.40, SD = 1.19) scenarios, and that moderate levels of involvement (M = 4.12, SD = 0.89) were rated as more likely than full levels of involvement (M = 3.03, SD = 1.33). However, there was no significant interaction effect, *F*(1, 75) = 0.14, *p* = .711.

#### Living-Well

At the Living-Well stage (Fig 2d), there were significant main effects of Level of Involvement, *F*(1,75) = 63.28, *p* <.001, n^2^_p_ = 0.46 95%CI [0.29, 0.58], and of Centrism, *F*(1,75) = 21.41, *p* <.001, n^2^_p_ = .22, 95%CI [0.08, 0.38]. This indicated that human-centric (M = 3.99, SD = 1.18) scenarios were rated as more likely than AI-centric (M = 3.41, SD = 1.14) scenarios, and that moderate levels of involvement (M = 4.26, SD = 0.75) were rated as more likely than full levels of involvement (M = 3.15, SD = 1.30). However, there was no significant interaction effect between the 2 variables, *F*(1, 75) = .00, *p* = .958.

#### Dying-Well

At the Living-Well stage (Fig 2e), there were significant main effects of Level of Involvement, *F*(1,75) = 53.29, *p* <.001, n^2^_p_ = 0.42 95%CI [0.25, 0.55], and of Centrism, *F*(1,75) = 27.24, *p* <.001, n^2^_p_ = 0.27, 95%CI [0.11, 0.42]. This indicated that moderate levels of involvement (M = 4.27, SD = 0.85) were rated as more likely than full levels of involvement (M = 3.37, SD = 1.27), and that Human-centric scenarios (M = 4.14, SD = 1.08) were deemed more likely than AI-centric scenarios (M = 3.49, SD = 1.17). However, no significant interaction effects were discovered, *F*(1, 75) = 2.68, *p* = .106.

### Influence of Dementia Knowledge and AI Literacy

To examine whether individual differences influenced perceptions of system acceptability and likelihood, additional analyses were conducted with variables that recorded dementia knowledge, dementia awareness, and AI literacy. Full statistical details are reported in the Supplementary Information, high-level summarises of outcomes are provided below.

#### Dementia Knowledge

Scores on the Dementia Knowledge Assessment Tool (DKAT) ranged from 3 to 19 (out of 21), with a mean of 12.97 (SD = 3.30), indicating that participants possessed moderate, foundational knowledge of dementia. Including DKAT as a covariate did not alter any of the primary findings for either perceived acceptability or likelihood. All main and interaction effects remained significant in the same direction as in the original analyses. This suggests that differences in dementia knowledge did not significantly influence participants’ evaluations of human- versus AI-centric systems or involvement levels across dementia care stages.

#### Dementia Attitudes

Scores on the Dementia Attitude Scale (DAS) ranged from 75 to 128 (out of 140), with a mean of 98.07 (SD = 11.99), reflecting generally positive attitudes toward people with dementia. In addition, 59.2% of participants reported personally knowing someone with dementia, underscoring the relevance and personal context of responses. When DAS scores were added as a covariate, no changes occurred in the significance or direction of the main or interaction effects for either acceptability or likelihood. This indicates that the observed effects were robust beyond general attitudinal differences, and that participants’ positivity toward dementia care did not bias their responses to the AI- versus human-centric scenarios.

#### AI Literacy

Scores on the Artificial Intelligence Literacy Scale (AILS) ranged from 24 to 78 (out of 94), with a mean of 59.75 (SD = 11.21), suggesting a moderate to high level of AI literacy among participants. Introducing AILS as a covariate likewise did not change the overall pattern of results. All main and two-way interaction effects remained significant, with only the three-way interaction for perceived likelihood (Stage × Centrism × Involvement) becoming non-significant. This indicates that while AI literacy may influence how participants conceptually interpret AI involvement, it did not systematically moderate or diminish the core relationships observed in the primary analyses.

Across all models, incorporating DKAT, DAS, and AILS scores as covariates did not alter the substantive outcomes. The main effects of stage, level of involvement, and centrism, as well as most interaction effects, were replicated and stable. The only exception was a reduction in the three-way interaction for perceived likelihood, which became non-significant when controlling for individual differences. Overall, these findings suggest that participants’ knowledge, attitudes, and literacy regarding dementia and AI did not meaningfully confound or moderate their evaluations of system acceptability or likelihood.

## Discussion

The purpose of this study was to investigate how the acceptability and adoption of AI-integrated dementia care are influenced by the stage of the care pathway, system centrism (AI versus human), and the level of involvement of the system in patients’ lives. Our findings demonstrate that public perceptions are highly context-dependent, with nuanced attitudes that shift as patients progress through the care pathway. We can infer from these findings that the acceptance of AI technology within dementia care is a multidimensional construct, shaped by a dynamic interplay of ethical, relational, and practical considerations rather than as a simple binary judgment of “yes” or “no.” This complexity highlights a central principle in digital health: technology integration is not merely a matter of capability but must align with human values, care contexts, and societal expectations.

A particularly salient result was the significant three-way interaction between Stage, Centrism, and Level of Involvement for acceptability. Attitudes toward AI versus human-centric systems were not fixed but evolved according to the patient’s progression through the dementia-care pathway. In the early Preventing-well and Diagnosing-well stages, higher acceptance ratings indicated a cautious openness to AI systems, even when these were designed for higher involvement. This finding suggests that AI technologies can be acceptable if framed appropriately, particularly as tools that enhance autonomy, support preventative measures, or facilitate early diagnosis. This observation is consistent with broader technology acceptance literature, which emphasises that perceived usefulness, transparency, and control are crucial determinants of adoption [33, 34]. For example, framing AI as an assistive partner rather than a replacement may help mitigate resistance and enhance user confidence.

As patients move into later stages (Supporting-well, Living-well, and Dying-well) the data indicate a pronounced preference for fully involved, human-centric systems. In these stages, acceptability shifts sharply away from AI-dominant scenarios, reflecting heightened concerns about trust, ethical responsibility, privacy, and dignity. The Dying-well stage underscores the ethical sensitivity of end-of-life care; participants rated highly involved AI systems as less acceptable, likely perceiving them as intrusive or diminishing human agency. Empathy on behalf of the participants may also play an important role in determining what is acceptable or unacceptable in end-of-life care [35], where scenarios that elicit more empathetic responses may cause participants to express more sensitivity and consideration when deciding what is acceptable. This aligns with prior research highlighting that acceptance of automation is contextually contingent and deeply entwined with perceived risk, moral responsibility, and the potential for loss of control [36]. Notably, the Supporting-well stage showed significant main effects for human centrism and full involvement, whereas in the Living-well and Dying-well stages, these preferences were revealed primarily through interaction effects, emphasising the compounded influence of multiple factors in high-stakes care contexts. This aligns with current technology acceptance theories such as the Unified Theory of Acceptance and Use of Technology (UTAUT2) model which notes that there are multiple factors affecting users’ intentions, which are then also moderated by other characteristics and contexts [33, 37]

Interestingly, moderately involved AI systems were consistently perceived as acceptable across all stages. This suggests that AI can occupy a supportive, collaborative role without triggering concerns over autonomy or ethical compromise. Such findings support the notion of adaptive autonomy in digital health design, where the degree of system involvement can be tailored to individual patient and caregiver needs. Moderately involved AI systems may function as decision-support tools, enhancing efficiency and care quality while maintaining a human-in-the-loop framework that is critical for sensitive and complex healthcare contexts. Examples of this may include risk assessment support derived from passive lifestyle monitoring by an AI system that can flag potential risk factors to clinicians who would then discuss modifications with the patient. In this way, the AI does not control any aspect of the patient’s life, nor does it remove any agency or intrude in any way.

When evaluating likelihood, the patterns were clearer and more consistent. Human-centric systems with moderate involvement were generally seen as the most probable scenario, particularly in later stages of the dementia-care pathway. This links closely to theories of risk perception such as the Risk as Feelings Theory [38] which suggests that scenarios that invoke more empathy during decision making, such as the later, more sensitive stages of dementia care, can override cognitive analysis and distort likelihood ratings. This divergence between perceived acceptability and likelihood highlights an important consideration: the public’s view on the adoption of AI may be malleable, influenced by factors such as perceived risk or personal innovativeness [39] as well as direct characteristics of the AI itself. Such findings have direct implications for digital health policy and implementation strategies, suggesting that any integration of AI must balance innovation with realistic expectations of technology readiness, professional norms, and societal trust.

From a translational perspective, these findings carry substantial implications for developers, clinicians, and policymakers. Technology design should prioritize adaptive, patient-centred approaches, incorporating flexibility in system involvement according to stage-specific needs. Human-in-the-loop models are particularly critical in later stages of care to safeguard autonomy, dignity, and relational trust. Policy frameworks should reinforce ethical safeguards, transparency, and human oversight while encouraging innovation in early diagnosis and preventative care. Moreover, the consistent acceptability of moderately involved AI indicates a practical pathway for implementation. AI may be integrated as a collaborative, augmented tool rather than a fully autonomous replacement, supporting decision-making and easing caregiver workload without compromising patient values. These insights align with broader principles in digital health, emphasising user-centred design, ethical deployment, and context-sensitive automation [40, 41].

Methodologically, the study leveraged vignettes to explore complex, hypothetical care scenarios, providing rich, nuanced data. However, this approach has inherent limitations. Participants responded to imagined scenarios rather than interacting with tangible systems, which may omit critical real-world factors such as usability, interface intuitiveness, system reliability, and affective experience. This limitation underscores the need for future studies to enhance ecological validity, for instance, by incorporating interactive prototypes or Wizard-of-Oz paradigms that simulate real-world interactions with AI systems [42]. Longitudinal designs would also be valuable, tracking changes in public attitudes as AI technologies evolve and as societal exposure to AI in healthcare increases.

Furthermore, the rapidly advancing nature of AI in digital health suggests that public perceptions are likely to shift over time. Cross-sectional snapshots capture contemporary attitudes but cannot account for temporal dynamics, changes in trust, or exposure-driven adaptation. Future research should therefore integrate longitudinal assessments to provide a more comprehensive understanding of technology acceptance trajectories, identifying factors that can positively or negatively influence adoption, trust, and perceived appropriateness over time.

In conclusion, this study demonstrates that public perceptions of AI-integrated dementia care are highly context-dependent and shaped by the interplay of care stage, system centrism, and level of involvement. Human-in-the-loop approaches are consistently preferred in later, more vulnerable stages, while moderately involved AI systems offer a broadly acceptable pathway across the care continuum. These findings carry important implications for technology design, policy development, and clinical practice. With our sample being made up of exclusively 55–64-year-olds, who will be the next generation to utilise technologies such as this, allows this study to highlight the necessity of adaptive, ethically grounded, and patient-centred approaches in digital health interventions, reinforcing that successful AI integration depends on trust, dignity, and the careful negotiation of human and technological roles. By acknowledging the nuanced perspectives of patients, carers, and the wider public, digital health stakeholders can ensure that AI-enhanced dementia care is not only technologically feasible but socially acceptable, ethically robust, and ultimately supportive of human agency across all stages of the care pathway.

## Acknowledgments

We would thank participants for their contribution to this research. This work was funded by Northumbria University.

## Author contributions

All authors conceptualised the study. T.O. developed the data collection materials and performed data analyses and drafted the manuscript. S.V and M.C provided feedback and revisions on the manuscript. All authors approved the content of the manuscript before submission.

## Data availability statement

Data collection materials and the data that support the findings of this study are freely available on the project OSF site at https://osf.io/wautg/.

